# Contactless Camera-Based Home Measurement of Pre-Attack Physiology in Migraine: A Longitudinal Feasibility Study

**DOI:** 10.64898/2026.09.13.26362930

**Authors:** Gertjan Van Gils

## Abstract

**Aim:** To develop and evaluate a contactless, camera-only method for measuring pre-attack physiology in migraine at home, and to assess its feasibility over four weeks.

**Methods:** In this longitudinal observational feasibility study, adults with a neurologist diagnosis of migraine used a browser-based, camera-only application on their own device three times daily for four weeks. The application derived heart rate and heart-rate variability from facial video, together with proxies for eyelid and mouth movement and blink rate, each referenced to a person-specific baseline. Measurements within twelve hours before a self-reported attack were compared with attack-free measurements using linear mixed models, and individual prediction was tested with leave-one-subject-out cross-validation. The complete analysis was independently re-implemented and reproduced.

**Results:** Nine of thirteen recruited participants completed the study and contributed 549 measurements, with high data completeness (547 reliable). Heart rate was higher in the twelve hours before an attack than in attack-free periods (66.0 versus 60.4 beats per minute), confirmed by a mixed model (plus 5.54 beats per minute) and unchanged after adjustment for stress; no other signal differed after correction. Individual-level prediction remained weak (best cross-validated area under the curve 0.58).

**Conclusion:** Contactless, camera-only measurement of pre-attack physiology at home is feasible and reproducible. A group-level rise in heart rate was demonstrable but did not translate into usable individual prediction, motivating a larger study with electrocardiographic validation.

## 1. Introduction

Migraine is a leading cause of disability in adults under 50 [1]. Acute treatment works best when taken early, and can even abort the headache when taken during the prodrome, before pain begins [2,3]; the ability to anticipate an attack could therefore improve treatment timing and quality of life. Yet the physiological changes that precede an attack are difficult to capture with everyday, unobtrusive tools, which limits their use outside specialist settings.

Two lines of evidence motivate a camera-based approach. First, changes in facial expression have been linked to migraine: Chen et al. reported that facial-expression changes in patients with migraine, namely increased eyelid tightening and reduced mouth movement, track headache pain, with eyelid tightening significantly correlated with pain intensity [4]. Second, cardiac autonomic function, reflected in both heart rate and its variability (HRV), is altered across the migraine cycle [5]. HRV can decline in the pre-migraine period, sometimes hours before an attack [6–7], and continuous ambulatory monitoring of haemodynamic variables, including heart rate, has been used to anticipate attacks, with per-patient models yielding useful forecast windows [8]. Whether mean heart rate itself, as distinct from its variability, changes measurably before an attack is, however, less well established, and is one of the questions this study addresses. Examining it is nonetheless well motivated, for two reasons. First, because heart rate is governed by the same autonomic system whose dysregulation is evident in migraine, a pre-attack change in heart rate is biologically plausible, so the hypothesis is grounded rather than speculative. Second, and importantly for a home-based tool, heart rate can be recovered far more reliably than HRV from an ordinary camera: HRV estimation requires many clean inter-beat intervals and is highly sensitive to noise over short recordings, whereas mean heart rate is comparatively stable [9]. A simple, dependably measurable signal that carried information about an impending attack would therefore be especially valuable for an accessible, at-home application. More broadly, mobile-health and wearable data combined with machine learning have been used to forecast migraine attacks [10–14]; however, reported performance remains modest and depends strongly on how the models are validated and which outcome is predicted, a point we return to when interpreting our own results.

For the present study we selected a set of signals that can be derived from an ordinary camera, supplemented by ambient data, and that have a physiological or empirical link to migraine. Heart rate and HRV index the autonomic function discussed above [5–8]; the AU7 proxy (eyelid tension) and the AU25 proxy (mouth opening) capture facial-expression changes associated with head pain [4]; spontaneous blink rate was recorded as an exploratory, non-invasive marker of central dopaminergic activity [15], a system implicated in the migraine premonitory phase; and barometric pressure was included as environmental context, because weather and pressure changes are among the most commonly reported migraine triggers [16].

Electrophysiological approaches, in particular electroencephalography (EEG), can detect neural changes that precede a migraine attack [17–20], but they require dedicated hardware: conventional recordings are largely confined to clinical or laboratory settings [17], and even the easy-to-use wearable EEG systems developed for home use still depend on a device worn on the head [18–19]. Body-worn sensor networks and wearables are more portable, yet also depend on hardware that must be worn and charged [8,10–11]. A measurement using only a camera the person already owns could substantially lower this barrier for continuous, real-world monitoring, albeit at the cost of greater signal noise. Remote photoplethysmography (rPPG) allows heart rate and, with caution, HRV to be estimated contactlessly from ordinary camera video [21–22], while facial-landmark tracking enables proxies for facial action units [23]. However, to our knowledge no previous study combines facial-action-unit proxies and rPPG-derived HRV in a single, purely camera-based system that uses a person-specific baseline together with longitudinal time-labelling of attacks.

This pilot study addresses that gap by developing and evaluating a contactless, camera-only measurement pipeline rather than by testing a clinical predictor. Its objectives are (i) to describe the measurement and signal-processing pipeline, which combines rPPG-derived heart rate and HRV with the AU7 and AU25 facial proxies, a person-specific baseline, and a transparent, independently reproduced analysis; (ii) to assess its feasibility, data completeness and usability in a four-week home setting; and (iii) as a demonstration of the pipeline, to examine whether the camera-derived signals change measurably in the hours before a migraine attack and to evaluate a person-specific predictive model. Given the exploratory nature and small sample of a pilot, the results are intended to be preliminary and to inform the design and sample size of a larger confirmatory study.

## 2. Materials and Methods

### 2.1. Study Design

This was an observational, longitudinal pilot study with repeated measures following an ecological momentary assessment design. The study was non-interventional: it involved no invasive procedures, no administration of medication, no clinical alarms and no involvement of health-care providers. The study was conducted under protocol MIG-PILOT-2026-001, which is available from the author on request. It was classified as low risk, and participants could withdraw at any time without giving a reason and without consequences. The study was not prospectively registered in a public registry. No patients or members of the public were involved in the design, conduct, or reporting of the study.

### 2.2. Participants

Eligible participants were adults aged 18–65 years with a clinical diagnosis of episodic or chronic migraine and at least one attack per month who owned a camera-equipped personal device. The sample size was determined by feasibility as a pilot rather than by a formal power calculation, and the study was not powered for individual-level prediction. Exclusion criteria were eye conditions liable to affect the eye-aspect-ratio (EAR) measurement, motor disorders limiting facial expression, pregnancy, and acute psychiatric disorders.

Age and sex were recorded at inclusion.

### 2.3. Measurement Application

Measurements were obtained with a standalone HyperText Markup Language application that runs locally in a web browser (Chrome or Safari) on a smartphone, tablet or laptop, without installation and without a server. No images or video were stored; only computed measurement values were retained. In line with MDCG 2019-11, the application was qualified as general-purpose software not falling under the Medical Device Regulation, providing no diagnosis, no treatment advice, no clinical alarm and no health-care-provider access. No commercial use was intended at this stage, and the application was made available only to study participants under the supervision of the principal investigator.

### 2.4. Measurement Protocol

Participants completed measurements three times daily (morning, midday and evening) for four weeks. Each measurement lasted 60 s and comprised three 20-s segments: (1) neutral gaze, for rPPG and HRV; (2) eyes held wide open, for the AU7 proxy; and (3) a closed-lip smile, for the AU25 proxy. Each session also collected self-reported sleep quality (0–10, morning only), pain on a 0–10 numeric rating scale, migraine phase, stress, medication and premonitory symptoms. At every subsequent measurement, participants indicated whether an attack had occurred since the previous one (no; yes, <6 h; yes, 6–12 h); this generated the time-label used for the predictive analysis. Following a screening and inclusion window (weeks 1–2), a person-specific baseline was established in week 3 (at least seven attack-free measurements over at least three days), with longitudinal measurement in weeks 4–6; the overall measurement period thus spanned approximately four weeks. Recruitment and data collection took place between June and July 2026.

### 2.5. Signal Processing

#### 2.5.1. Heart Rate (via rPPG)

Heart rate was derived from rPPG using the chrominance method [21]. Per-frame red-green-blue values were extracted from six cheek points located with MediaPipe Face Mesh. After a 5-s calibration on a white reference, the pulse signal was computed as 3×(Rn/tot) − 2×(Gn/tot), with tot = Rn + Gn + Bn, and heart rate was obtained by a discrete Fourier transform with a Hanning window over 0.67–3.0 Hz (40–180 bpm) [22].

#### 2.5.2. Heart-Rate Variability

HRV was quantified by two time-domain parameters: the standard deviation of NN intervals (SDNN) and the root mean square of successive differences (RMSSD). These were chosen because they are the standardised time-domain measures for which normative reference values are available, and because time-domain measures are more reliably estimated from short recordings than frequency-domain measures, which require longer, stable segments for reliable spectral analysis [9]. SDNN was computed from the RR intervals (Bessel correction, n−1; filter 400–1500 ms) and RMSSD following the Task Force standards [24]. RR peaks were detected with a signal threshold (0.3σ), a refractory period (≥200 ms between peaks) and an outlier filter removing RR intervals deviating more than 50% from the median. Each measurement spans only ∼22 s (yielding approximately 20–28 RR intervals), whereas the clinical standard requires at least 5 min [24].

#### 2.5.3. Facial Action Unit Proxies

Facial landmarks were obtained with MediaPipe Face Mesh. The AU7 proxy (eyelid tension) used the six-point EAR, computed as (v1+v2+v3)/(3×h) [23] and was measured in segment 2. The AU25 proxy (mouth opening) used the ratio of vertical to horizontal mouth distance and was measured in segment 1. Blink frequency was counted as EAR drops below 0.13 within the neutral segment and expressed per minute. These are geometric approximations, the EAR for the AU7 eyelid-tension proxy and the vertical-to-horizontal mouth ratio for the AU25 mouth-opening proxy, operationalised as action-unit proxies; they do not constitute certified Facial Action Coding System (FACS) coding and are not identical to the specific action units used in reference FACS studies [4] (for example, the AU25 proxy approximates mouth opening rather than the mouth-stretch action, AU27).

#### 2.5.4. Ambient Pressure and Data-Quality Indicators

Barometric pressure (absolute value and change relative to the previous measurement) was obtained from the Open-Meteo application programming interface using the device GPS location. Data-quality indicators recorded per measurement were rPPG success (ok, 0/1), the number of rPPG frames (≥600 recommended for reliable HRV) and the proportion of frames with successful face detection. Failed measurements produced null values, which were not imputed.

### 2.6. Person-Specific Baseline and In-App Risk Estimate

After at least seven attack-free measurements spread over at least three days, a person-specific baseline was computed for each signal. The application displayed an experimental risk estimate (0–100%) defined heuristically as Score = 50 − zHRV×15 + zHR×10 + zAU7×12 − zAU25×8 + zRMSSD×5 (bounded at ±2σ). This estimate was explicitly labelled as an experimental model output.

### 2.7. Statistical Analysis

Differences in each signal between pre-attack (within 12 h) and attack-free measurements were first examined with the Mann–Whitney U test, with Benjamini–Hochberg false-discovery-rate (FDR) correction across the signals tested. Two complementary definitions of attack proximity were used: the migraine phase recorded at each measurement (attack-free, phase 0, versus during an attack, phase 2), which underlies the attack-free-versus-attack comparison and the per-participant attack counts in Table 1; and a pre-attack label derived from the prospective feedback (whether an attack followed within 12 h), which underlies the primary pre-attack analysis, the linear mixed model and the predictive validation. Because these are defined differently, their measurement counts differ. Because this test treats the repeated measurements from each participant as independent, the primary analysis used linear mixed models (restricted maximum likelihood, random intercept per participant; signal ∼ pre-attack + (1|participant)) to account for repeated measurements within participants. This model was fitted separately for each signal, and the resulting p-values were FDR-corrected across signals. Predictive performance was evaluated with a logistic-regression model under leave-one-subject-out (LOSO) cross-validation, with the baseline, normalisation and decision threshold determined per fold on the training data to avoid leakage. No hyperparameter tuning was performed; the model was fitted with default settings. A held-out participant contributed a fold only if they had at least one pre-attack-labelled and one attack-free-labelled measurement; participants lacking either class were not evaluable. Discrimination was summarised with the area under the receiver-operating-characteristic curve (ROC-AUC) and the precision–recall AUC (PR-AUC) referenced to the sample prevalence; positive and negative predictive values (PPV/NPV) were reweighted to the expected base rate using Bayes’ rule. Model calibration was not formally assessed, given the small sample size and the exploratory aim of the individual-level analysis. Univariate Pearson correlations were used only for rough feature exploration and, because they ignore the repeated-measures structure, are not interpreted as model importance, since feature-attribution and explainability estimates are themselves unstable when predictors are correlated [25]. The number of events per predictor was computed for each model as an indicator of overfitting risk [26]. This observational study is reported in accordance with the STROBE guideline for observational studies (checklist provided in Document S2); the predictive-modelling component additionally follows the TRIPOD+AI reporting guideline [27] (checklist provided in Document S3).

**Table 1.** Data yield and baseline status per participant.

| Participant | Measurements | Reliable (%) | Attack-free | Attacks | Feedback (%) |
| --- | --- | --- | --- | --- | --- |
| P1 | 123 | 123 (100) | 37 | 11 | 94 |
| P2 | 84 | 82 (98) | 49 | 16 | 70 |
| P3 | 80 | 80 (100) | 78 | 1 | 65 |
| P4 | 80 | 80 (100) | 48 | 13 | 52 |
| P5 | 68 | 68 (100) | 50 | 0 | 12 |
| P6 | 61 | 61 (100) | 35 | 21 | 69 |
| P7 | 21 | 21 (100) | 12 | 2 | 81 |
| P8 | 18 | 18 (100) | 11 | 6 | 67 |
| P9 | 14 | 14 (100) | 11 | 0 | 79 |
| Total | 549 | 547 (99.6) | 331 | 70 | 65 |

Attacks: measurements recorded during an attack (migraine phase 2).

All analyses reported here were computed by the study application and were independently re-implemented in Python 3.12 to verify their correctness. The Mann–Whitney U tests were reproduced with SciPy, the linear mixed models (restricted maximum likelihood, random intercept per participant) with statsmodels, and the logistic-regression and leave-one-subject-out ROC/PR analyses with scikit-learn. The pre-attack labelling and the per-participant 24-h barometric-pressure change were applied identically in both pipelines. Descriptive statistics, the non-parametric group comparisons, the mixed-model coefficients and variance decomposition (ICC), the time-window means and the cross-validated discrimination reproduced to three to four decimal places, confirming that the in-application computations agree with these established reference libraries. The independent reference implementation is provided as Code S1.

## 3. Results

### 3.1. Participant Flow and Feasibility

Thirteen eligible participants (2 men, 11 women) were recruited. Four women did not complete the study: one could not use the application because facial recognition failed to capture all required parameters, and three did not meet the criterion for establishing a person-specific baseline by week 3, with fewer than seven attack-free measurements recorded across at least three days. The remaining 9 participants were included in the analysis. Two participants had episodic and seven had chronic migraine according to the treating neurologist’s diagnosis; the number of attacks logged during the study window (Table 1) varied and did not always reflect this classification, as the app captured only a limited period with variable adherence.

The 9 analysed participants contributed 549 measurements in total (median 68 per participant, range 14–123). Of these, 547 (99.6%) were technically reliable (ok = 1) and 2 failed. In total, 70 measurements were recorded during an attack, 359 carried self-reported feedback, and 102 were labelled as occurring within 12 h before an attack. Every participant reached a valid person-specific baseline of at least seven attack-free measurements. Adherence varied widely between participants (Table 1). The analysed participants had a mean age of 38.6 ± 11.4 years (range 26–56).

Data completeness was high for the camera-derived signals: heart rate 100%, the AU7 and AU25 proxies 100%, blink rate 100%, and HRV (SDNN) and HRV (RMSSD) 98% (null when too few RR intervals were available). Barometric pressure was 82% complete (GPS declined or no internet for the remainder) and sleep, which was reported once daily (each morning, about the previous night) and therefore applies to only one of the three daily sessions by design, was 35% complete (Table 2).

**Table 2.** Data completeness per measured variable across all 549 measurements.

| Variable | Non-null | Null | Complete (%) |
| --- | --- | --- | --- |
| Heart rate | 547 | 2 | 100 |
| HRV (SDNN) | 540 | 9 | 98 |
| HRV (RMSSD) | 540 | 9 | 98 |
| AU7 proxy (EAR) | 547 | 2 | 100 |
| Blink rate | 549 | 0 | 100 |
| AU25 proxy (mouth) | 547 | 2 | 100 |
| Barometric pressure | 452 | 97 | 82 |
| Sleep | 190 | 359 | 35 |

Null values indicate a failed or unavailable measurement and were not imputed. Sleep was collected only in the morning session. AU = action unit; EAR = eye-aspect ratio; HRV (RMSSD) = heart-rate variability (root mean square of successive differences); HRV (SDNN) = heart-rate variability (standard deviation of NN intervals).

### 3.2. Signal Changes Before Attacks

The primary question was whether signals differ in the hours before an attack. When pre-attack measurements with a valid heart rate (within 12 h; n = 96) were compared with attack-free measurements (n = 262), heart rate was significantly higher before an attack (66.0 ± 19.0 vs. 60.4 ± 18.7 bpm; Mann–Whitney U test, p = 0.004, significant after FDR correction), whereas HRV (SDNN and RMSSD), the AU7 and AU25 proxies and blink rate did not differ significantly (Table 3, Figure 1). All pre-attack and no-attack measurements had a valid heart rate, so the descriptive comparison, the heart-rate–based mixed model and the cross-validation use the same groups (n = 358 overall).

**Figure 1.**
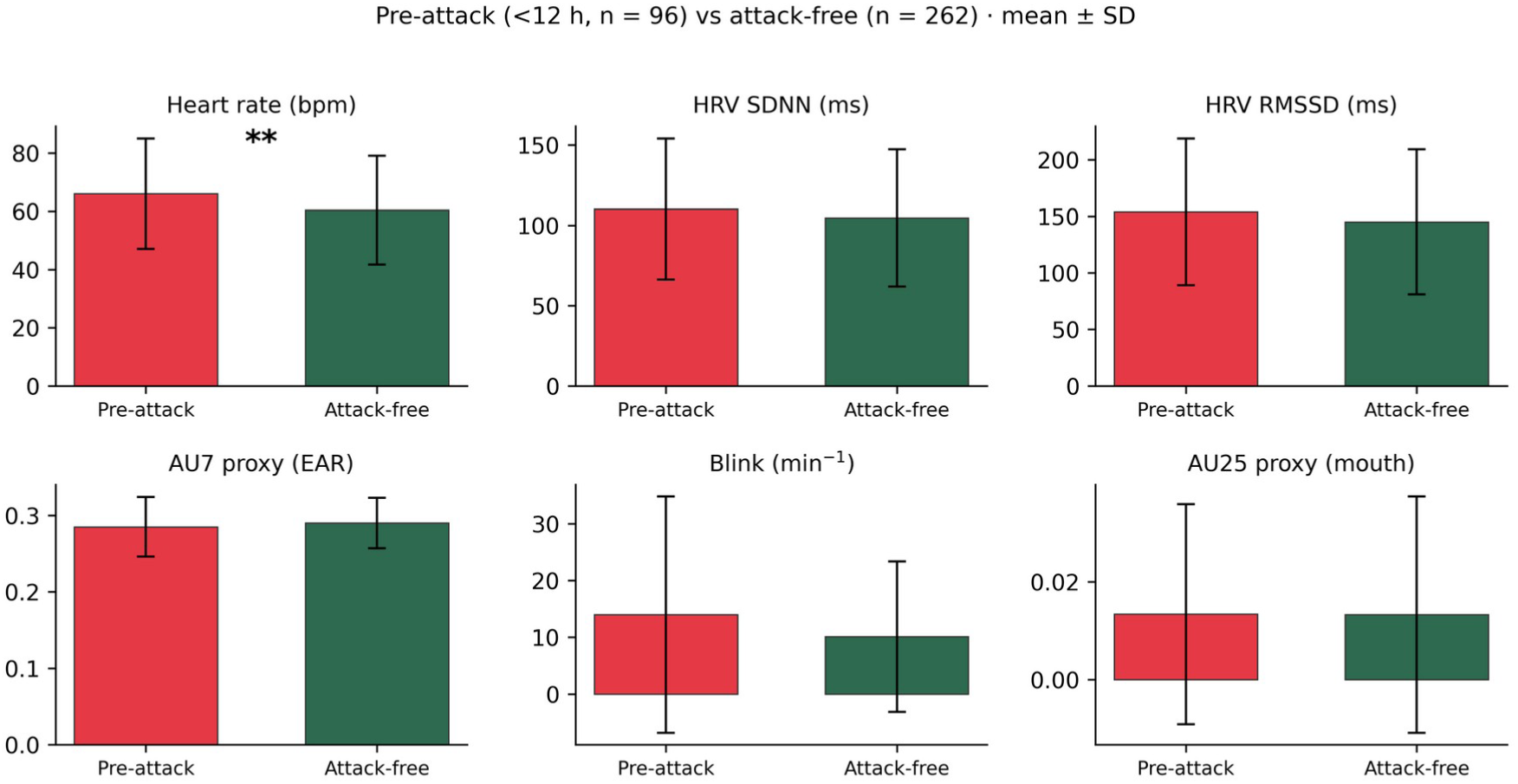
Signal values in pre-attack (<12 h) versus attack-free measurements. Bars show mean ± SD; heart rate was significantly higher before attacks (** p = 0.004, FDR-corrected), whereas HRV (SDNN) and HRV (RMSSD) did not differ. bpm = beats per minute; FDR = false discovery rate; HRV (RMSSD) = heart-rate variability (root mean square of successive differences); HRV (SDNN) = heart-rate variability (standard deviation of NN intervals); ms = milliseconds; SD = standard deviation.

**Table 3.** Signal values in the 12 h before an attack versus attack-free measurements.

| Signal | Pre-attack (mean ± SD) | No-attack (mean ± SD) | p | Direction |
| --- | --- | --- | --- | --- |
| Heart rate (bpm) | 66.03 ± 18.98 | 60.40 ± 18.65 | 0.004 * | higher pre-attack |
| HRV (SDNN) (ms) | 110.17 ± 43.90 | 104.56 ± 42.72 | 0.275 | higher (ns) |
| HRV (RMSSD) (ms) | 154.00 ± 64.87 | 145.18 ± 64.15 | 0.283 | higher (ns) |
| AU7 proxy (EAR) | 0.28 ± 0.04 | 0.29 ± 0.03 | 0.540 | lower (ns) |
| Blink (min<sup>-1</sup>) | 13.97 ± 20.81 | 10.09 ± 13.25 | 0.062 | higher (ns) |
| AU25 proxy (mouth) | 0.01 ± 0.02 | 0.01 ± 0.02 | 0.298 | higher (ns) |

Pre-attack $n = 96$ , no-attack $n = 262$ ; Mann–Whitney $U$ test. \* significant after false-discovery-rate correction. AU = action unit; bpm = beats per minute; EAR = eye-aspect ratio; HRV (RMSSD) = heart-rate variability (root mean square of successive differences); HRV (SDNN) = heart-rate variability (standard deviation of NN intervals); ms = milliseconds; ns = not significant; SD = standard deviation.

The linear mixed model confirmed this effect: heart rate was on average 5.54 bpm higher before an attack (95% CI 1.33–9.75; p = 0.010; n = 358 measurements from 9 participants; intraclass correlation coefficient (ICC) 0.22, indicating that a mixed model was warranted). The effect persisted after adjustment for concurrent self-reported stress (β = +5.58 bpm, p = 0.012), while stress itself was non-significant, indicating that the pre-attack rise was not explained by momentary stress. Applying the same model to each signal confirmed heart rate as the only measure that remained significant after correction for multiple comparisons: HRV (SDNN) (β = +1.86 ms; p = 0.67), HRV (RMSSD) (β = +3.41 ms; p = 0.62), the AU7 proxy (β = −0.001; p = 0.87) and blink rate (β = +2.08 min⁻¹; p = 0.24) did not differ, and the AU25 proxy showed no significant change (β = −0.002; p = 0.35).

Across pre-attack time windows, mean heart rate increased as the attack approached, from 58.8 bpm at 12–24 h to 64.4 bpm at 6–12 h and 67.2 bpm at 0–6 h (Figure 2). When active attacks (phase 2) were compared with attack-free periods, the AU7 proxy was slightly but significantly lower (0.29 vs. 0.28; p = 0.014) and the AU25 proxy slightly higher (p = 0.030), while heart rate and HRV did not differ.

**Figure 2.**
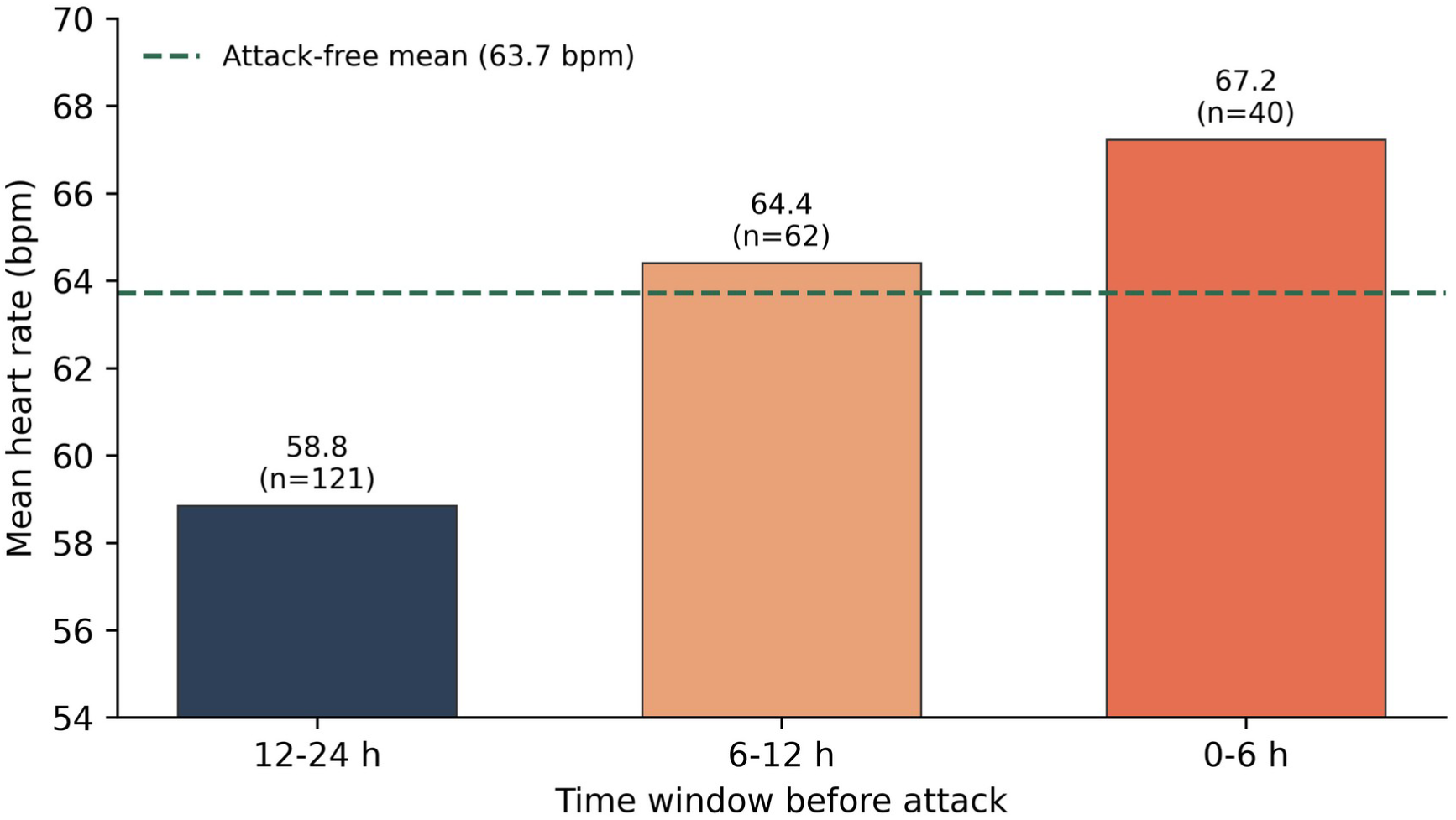
Mean heart rate by time window before an attack, showing a progressive rise as the attack approaches relative to the attack-free mean (dashed line). bpm = beats per minute.

### 3.3. Predictive Performance

Predictive performance was evaluated with LOSO cross-validation across six models (pre-attack n = 96, no-attack n = 262; Table 4). A univariate heart-rate model reached an in-sample AUC of 0.599. The best out-of-sample model (five signals plus barometric pressure) achieved a LOSO-AUC of 0.579 ± 0.220 (range 0.20–0.93 across folds; in-sample 0.630; Figure 3). At an assumed pre-attack prevalence of 15% in the target population (a modelling assumption for illustration, not the 27% proportion of pre-attack measurements in this attack-enriched sample), this corresponded to a sensitivity of 37%, a specificity of 67%, a PPV of 17% and an NPV of 86% (true positives 29, false positives 59, true negatives 122, false negatives 50). The PR-AUC (0.294) only marginally exceeded the prevalence baseline (0.268), and adding barometric pressure yielded a small LOSO-AUC gain over the five-signal model (0.530 to 0.579; ΔAUC ≈ 0.05); the model nonetheless remained well below a clinically useful level.

**Figure 3.**
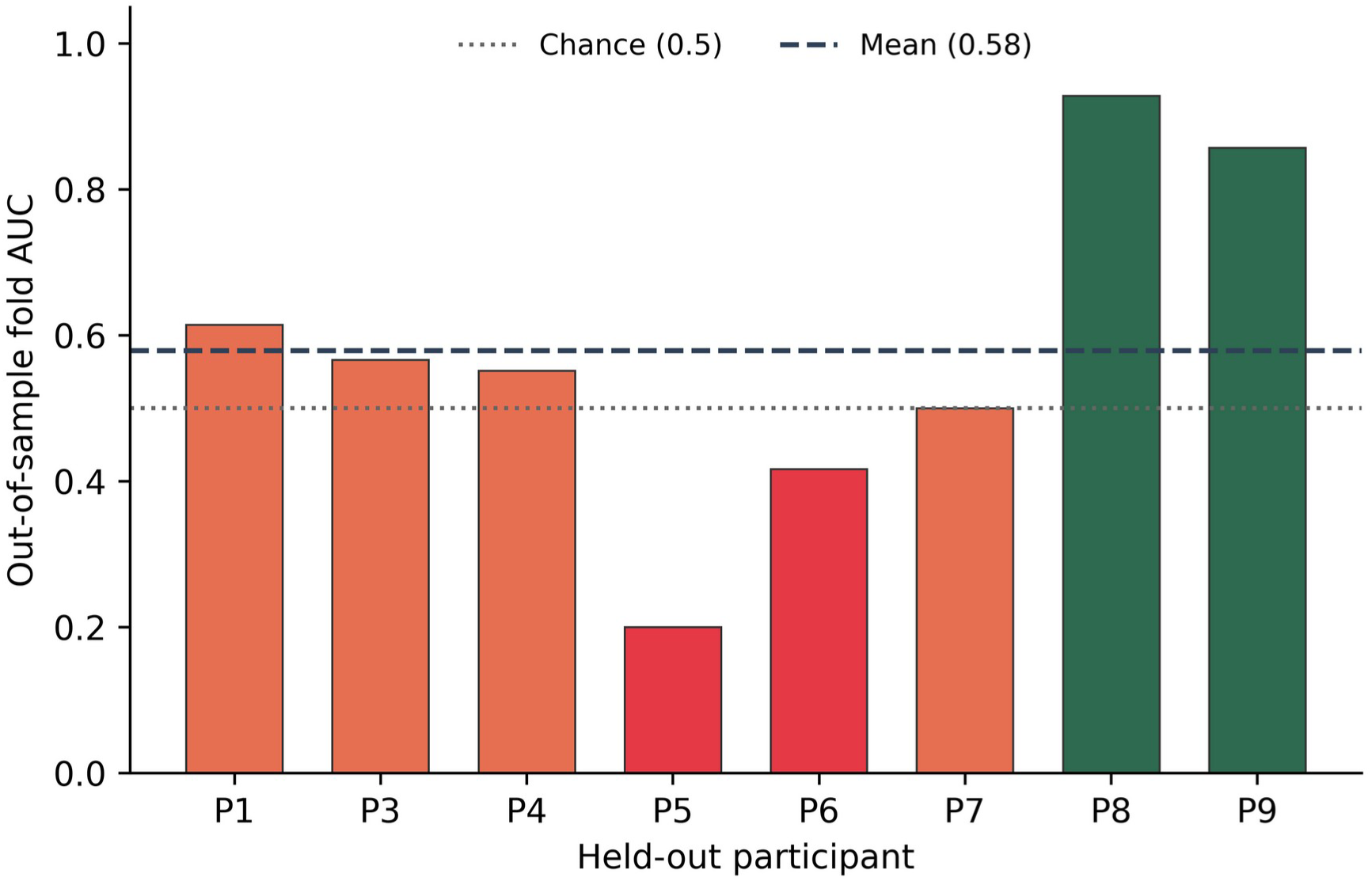
Out-of-sample fold AUCs from LOSO cross-validation of the best model, per held-out participant, with the chance level (0.5) and the mean (0.58) indicated. One participant (P2) lacked sufficient barometric-pressure data for the best model and could therefore not form a valid held-out fold (8 of 9 participants shown). AUC = area under the receiver-operating-characteristic curve; LOSO = leave-one-subject-out.

**Table 4.** LOSO cross-validated performance of the candidate predictive models.

| Model | Predictors | EPV | In-sample | LOSO-AUC | Sens. (%) | Spec. (%) | PPV (%) | PR-AUC |
| --- | --- | --- | --- | --- | --- | --- | --- | --- |

|  |  |  | **AUC** |  |  |  |  |  |
| --- | --- | --- | --- | --- | --- | --- | --- | --- |
| Univariate | HR | 96 | 0.599 | — | — | — | — | — |
| Multivariable (3) | HR + HRV (SDNN) + HRV (RMSSD) | 32 | 0.604 | 0.590 | 52 | 48 | 15 | 0.271 |
| Multivariable (5) | HR + HRV (SDNN) + HRV (RMSSD) + AU7 + AU25 | 19 | 0.615 | 0.530 | 57 | 45 | 16 | 0.263 |
| Δ-normalised (3) | HR + HRV (SDNN) + HRV (RMSSD), $\Delta$ vs. baseline | 32 | 0.611 | — | — | — | — | — |
| Multivariable (5)+pressure | HR + HRV (SDNN) + HRV (RMSSD) + AU7 + AU25 + pressure | 16 | 0.630 | 0.579 | 37 | 67 | 17 | 0.294 |
| Multivariable (5)+ΔP24 | HR + HRV (SDNN) + HRV (RMSSD) + AU7 + AU25 + Δpressure (24 h) | 16 | 0.625 | 0.517 | 32 | 60 | 13 | 0.300 |

*The five-signal-plus-pressure model (highlighted in the text) is the best out-of-sample model. AU = action unit; AUC = area under the receiver-operating-characteristic curve; EPV = pre-attack events per predictor; HR = heart rate; HRV (RMSSD) = heart-rate variability (root mean square of successive differences); HRV (SDNN) = heart-rate variability (standard deviation of NN intervals); LOSO = leave-one-subject-out; PPV = positive predictive value; PR-AUC = precision–recall AUC; Sens. = sensitivity; Spec. = specificity.*

Events per predictor were low (16–32 for the multivariable models), and all six models are reported to limit selection optimism. Out-of-sample discrimination below the commonly cited 0.70 threshold for clinical usefulness [28] indicates limited value at the level of an individual measurement, even though the group-level heart-rate effect is statistically robust; the association is not yet a usable individual predictor.

### 3.4. Exploratory Analyses

The most frequently reported premonitory symptoms were fatigue, neck stiffness and sound sensitivity, followed by nausea, cravings and aura. Per-participant comparisons of HRV (SDNN and RMSSD) between attack and attack-free measurements were heterogeneous in direction, with several participants showing higher rather than lower values around attacks; these exploratory analyses should be interpreted with caution given the short (∼22 s) recordings.

## 4. Discussion

This pilot developed and evaluated a contactless, camera-only measurement pipeline that combines facial-action-unit proxies and rPPG-derived heart rate and HRV with a person-specific baseline and an independently reproduced analysis, and tested its feasibility for longitudinal home monitoring in migraine. The primary contribution is the pipeline itself; the physiological results serve to demonstrate it. Three main findings emerged. The approach was feasible: 9 participants collected 549 measurements with high data completeness for the camera-derived signals. Heart rate was consistently elevated in the hours before an attack at the group level: it was significant in both the non-parametric comparison and a linear mixed model (+5.54 bpm; p = 0.010), robust to stress adjustment, and increased as the attack approached. However, this group-level signal did not translate into useful individual-level prediction, with the best LOSO model reaching an AUC of only 0.58.

These results are partly consistent with prior work. The pre-attack rise in heart rate aligns with reported autonomic involvement in the prodromal phase [6,8], and the small changes in the AU7 and AU25 proxies during attacks echo the association between facial expression and migraine pain reported by Chen et al. [4]. Whereas facial-expression changes and HRV alterations in migraine are relatively well documented [4–6], autonomic work has centred on HRV, and a reproducible pre-ictal rise in mean heart rate itself has been reported less often; demonstrating one here at the group level, holding after FDR correction and adjustment for stress and increasing towards attack onset, is a notable and novel result of this feasibility pilot. In contrast, HRV did not fall before attacks as hypothesised, and per-participant comparisons were mixed: several participants, including the one with the largest and most complete dataset, showed higher rather than lower HRV around attacks.

A likely explanation lies in the HRV measurement itself. The observed HRV (SDNN) values (per-participant attack-free means of roughly 60–200 ms, pooled mean ≈ 110 ms) and HRV (RMSSD) values (per-participant means ≈ 75–260 ms, pooled ≈ 151 ms) are physiologically implausible: in healthy adults, short-term resting SDNN is typically around 50 ms (rarely above ∼90 ms) and RMSSD around 40 ms (rarely above ∼75 ms) [29]. Such inflated values indicate that the HRV (SDNN) and HRV (RMSSD) estimates are dominated by measurement artefact rather than true autonomic variability. This is an expected consequence of deriving RR intervals from ∼22-s camera-based recordings, which yield only about 20–28 intervals and are sensitive to spurious or missed pulse peaks, since a single misdetected beat disproportionately inflates both indices. The HRV-based results should therefore be treated as unreliable and exploratory, and this artefact plausibly explains why HRV did not behave as hypothesised.

These predictive results should be read against the wider literature with care, following critical-appraisal principles for artificial intelligence in the headache field [30]. Higher AUCs are reported for migraine forecasting [10–14], but often under more favourable conditions: an in-sample or random hold-out split rather than subject-wise (LOSO) validation; an easier task (forecasting the next headache day from diaries rather than detecting a pre-attack state per measurement); and ROC-AUC rather than a metric suited to imbalanced data. Two distinctions matter. Out-of-sample versus in-sample: our five-signal model rose from a LOSO-AUC of 0.53 to an in-sample 0.61, the classic signature of overfitting. ROC-AUC versus PR-AUC: because attacks are rare, ROC-AUC can look respectable while PR-AUC, read against prevalence, reflects the precision a user would actually experience.

Against this background, our results match the most comparable work rather than being an outlier. Reported headline AUCs are often driven by high specificity on the many headache-free days: Stubberud et al. reported an AUROC of 0.62 whose best model had zero sensitivity [10], and Faisal et al. a time-series AUC of 0.84 with only 0.39 sensitivity [11]. Most directly, Holsteen et al. (178 participants, 1870 attacks) reported a within-person C-statistic of 0.56 and concluded that attacks were not predictable from self-report and triggers [12], close to our best LOSO-AUC of 0.58; stress-based forecasting (AUC 0.65) [13] and the more recent TRACE model (AUROC 0.66–0.67) [14] were similarly modest. Our PR-AUC (0.29) shows minimal lift over the positive-class prevalence (≈ 0.27); the apparent doubling of a 0.13 baseline is illusory, since 0.13 is the rarer prevalence of attack measurements among all measurements, an easier denominator that would overstate performance. Like the best comparable models, ours does not yet separate pre-attack from attack-free states at a clinically useful level [31].

### 4.1. Strengths and Limitations

The study has several strengths. Measurement was entirely contactless and image-free: no video or images were stored, the application ran locally on the participant’s own device without a server, and only computed values were retained, which lowers the barrier to participation and is favourable from a data-protection standpoint. Adherence and data completeness were high for the camera-derived signals (heart rate and the facial proxies ≥ 99% complete). The analysis was deliberately conservative and transparent: a person-specific baseline was used, predictive performance was estimated with leakage-free LOSO cross-validation, positive and negative predictive values were reweighted to a realistic base rate, PR-AUC was reported against prevalence, all six candidate models were reported rather than only the best, and repeated measurements were handled with a linear mixed model (ICC 0.22) with an explicit check for stress as a confounder. Within these safeguards, the group-level pre-attack rise in heart rate was statistically clear.

Several limitations temper interpretation. As a pilot, the sample is small and events per predictor are low (16–32), so predictive estimates are fragile, as the wide fold-level spread (0.20–0.93) shows. The sample was not demographically representative, so subgroup and fairness analyses (sex, age, and skin tone, the last particularly relevant for rPPG) were not possible, and generalisability to these groups is unknown. The HRV (SDNN) and HRV (RMSSD) indices are dominated by artefact from ∼22-s recordings and are not valid autonomic measures. The camera-derived heart rate was not validated against a simultaneous reference (electrocardiography, ECG, or pulse oximetry) in this home setting, although the underlying chrominance method has been validated previously [21]; because the pre-attack comparison was within-modality, a bias consistent within a participant largely cancels, but a state-dependent bias (e.g. restlessness or facial flushing before an attack) cannot be excluded, making concurrent reference validation a priority. This association-based, feasibility-oriented use of rPPG features follows an approach taken in recent work [32]. The facial measures are proxies, not certified FACS coding, and their dependence on lighting and reliable face detection made one participant unmeasurable. Attack labels were self-reported without clinical ground truth; barometric pressure was missing for 18% of measurements; sleep was reported only once daily, in the morning; and other acute drivers of heart rate (physical activity, caffeine, posture) were not recorded. Between-person heterogeneity was marked, with signals moving in opposite directions across participants, undermining a single universal model, and six models were compared on the same data, leaving some selection optimism. Most importantly, a group-level association is not a usable individual predictor: at the level of a single measurement the current models perform close to chance.

### 4.2. Implications and Future Directions

The findings identify a clear group-level pre-attack heart-rate signal while showing that individual prediction is not yet achievable with these signals alone. This modest individual-level performance is comparable to that of far more data-intensive approaches based on daily diaries and trigger reporting [10–14], suggesting that the limiting factor is not the richness of the data but the marked between-person heterogeneity of migraine; a small set of camera-derived signals is therefore a reasonable starting point, and the priority is to model each person individually rather than to fit a single generalised model, consistent with a recent review concluding that individualised models outperform generalised ones [33]. They motivate a larger confirmatory study (target n ≥ 50) with ECG validation, which would simultaneously provide a clinical ground truth and replace the unreliable camera-derived HRV with a valid measure. Future work should prespecify the analysis, use nested cross-validation for model selection, and move from a single universal model towards personalised, per-participant delta-feature models that compare each measurement with the individual’s own baseline, since the heterogeneity observed here suggests that individual signals may be stronger than pooled ones. Longer recordings, or dropping HRV in favour of the more reliably measured heart rate and facial signals, are concrete options to improve feature quality.

## 5. Conclusions

In this longitudinal pilot, a contactless, camera-only application capturing facial-action-unit proxies and rPPG-derived heart rate and HRV with a person-specific baseline was feasible in a home setting and achieved high data completeness. Heart rate was reproducibly elevated in the hours before a migraine attack at the group level (linear mixed model +5.54 bpm, p = 0.010), but this signal did not translate into reliable individual-level prediction (best LOSO-AUC 0.58). The findings are preliminary and not intended for clinical use; they provide a clear basis and effect estimates for a larger confirmatory study with ECG validation; such a study should also validate the camera-derived heart-rate variability against a reference standard, or use longer recordings, given the artefactually high variability observed here [24,29].

## Key Findings

- Contactless, camera-only measurement of pre-attack physiology at home was feasible over four weeks, with high data completeness.
- Heart rate rose in the hours before an attack at the group level, but individual-level prediction was not yet usable.
- A reproducible, image-free method provides a basis for larger, electrocardiography-validated studies.

## Declarations

## Data Availability

The data produced in the present study are available upon reasonable request to the corresponding author. The data are pseudonymised, camera-derived numerical measurements; individual-level data are not publicly available owing to privacy and data-protection (GDPR) restrictions.

## ACKNOWLEDGEMENTS

During the preparation of this work, the author used Claude (Anthropic) to assist with language editing and drafting of the text, with developing the code of the browser-based camera measurement application (dashboard) used for data collection, and with implementing and independently cross-checking the statistical analysis code reported here. The author reviewed and edited all AI-generated output and takes full responsibility for the content of the publication.

## ETHICAL CONSIDERATIONS

The study was conducted in accordance with the Declaration of Helsinki and approved by the accredited Ethics Committee of the ZAS hospital network (Ziekenhuis aan de Stroom, Antwerp, Belgium; participating sites: ZAS Cadix, ZAS Elisabeth, ZAS Erasmus, ZAS Hoge Beuken, ZAS Joostens, ZAS Middelheim, ZAS Palfijn, ZAS Paola and ZAS UKJA), approval number 009, on 10 June 2026 (protocol MIG-PILOT-2026-001). The application was qualified as non-MDR software under MDCG 2019-11. The study was not registered in a public trial registry.

## CONSENT TO PARTICIPATE

Informed consent was obtained from all participants involved in the study.

## CONSENT FOR PUBLISHING

The author agrees that, if accepted, the article will be published in Cephalalgia Reports.

## AUTHOR CONTRIBUTIONS

Gertjan Van Gils: Conceptualization; Methodology; Software; Formal analysis; Investigation; Data curation; Writing – original draft; Writing – review & editing; Visualization. The author has read and approved the final manuscript.

## DATA AVAILABILITY STATEMENT

Only pseudonymised, computed measurement values were retained (participants were identified solely by codes such as P1, P2); no images or video were recorded. Data were stored locally on participants’ own devices (browser localStorage) with no automatic server transfer, in accordance with the GDPR. The data are available from the corresponding author on reasonable request.

## PATIENT AND PUBLIC INVOLVEMENT

There was no patient or public involvement in the design, conduct, or reporting of this study.

## OPEN PRACTICES

This manuscript has not been posted on a preprint server.

## SUPPLEMENTAL MATERIAL

Document S1 (technical description of the measurement and signal-processing pipeline); Code S1 (analysis code); Document S2 (completed STROBE checklist); Document S3 (completed TRIPOD+AI checklist).

## References

1. Steiner TJ, Stovner LJ, Jensen R, Uluduz D, Katsarava Z. Migraine remains second among the world’s causes of disability, and first among young women: findings from GBD2019. J. Headache Pain 2020; 21: 137.

2. Goadsby PJ, Zanchin G, Geraud G, de Klippel N, Diaz-Insa S, Göbel H, et al. Early vs. non-early intervention in acute migraine—‘Act when Mild (AwM)’. A double-blind, placebo-controlled trial of almotriptan. Cephalalgia 2008; 28: 383–391.

3. Dodick DW, Goadsby PJ, Schwedt TJ, Lipton RB, Liu C, Lu K, et al. Ubrogepant for the treatment of migraine attacks during the prodrome: a phase 3, multicentre, randomised, double-blind, placebo-controlled, crossover trial in the USA. Lancet 2023; 402: 2307– 2316.

4. Chen WT, Hsiao FJ, Coppola G, Wang SJ. Decoding pain through facial expressions: a study of patients with migraine. J. Headache Pain 2024; 25: 33.

5. Zhang L, Qiu S, Zhao C, Wang P, Yu S. Heart Rate Variability Analysis in Episodic Migraine: A Cross-Sectional Study. Front. Neurol. 2021; 12: 647092.

6. Kapustynska V, Abromavičius V, Serackis A, Paulikas Š, Ryliškienė K, Andruškevičius S. Machine Learning and Wearable Technology: Monitoring Changes in Biomedical Signal Patterns during Pre-Migraine Nights. Healthcare 2024; 12: 1701.

7. Jankevičiūtė R, Kapustynska V, Andruškevičius S, Ryliškienė K, Abromavičius V. Heart rate variability as a predictor of migraine: Sleep-time data analysis of pre-migraine nights. Technol. Health Care 2026. 10.1177/09287329251412968.

8. Pagán J, De Orbe MI, Gago A, Sobrado M, Risco-Martín JL, Vivancos Mora J, et al. Robust and Accurate Modeling Approaches for Migraine Per-Patient Prediction from Ambulatory Data. Sensors 2015; 15: 15419–15442.

9. Munoz ML, van Roon A, Riese H, Thio C, Oostenbroek E, Westrik I, et al. Validity of (Ultra-)Short Recordings for Heart Rate Variability Measurements. PLoS ONE 2015; 10: e0138921.

10. Stubberud A, Ingvaldsen SH, Brenner E, Winnberg I, Olsen A, Gravdahl GB, et al. Forecasting migraine with machine learning based on mobile phone diary and wearable data. Cephalalgia 2023; 43: 3331024231169244.

11. Faisal F, Poole AC, Danelakis A, Wergeland T, Bjørk M, Kristoffersen ES, et al. Forecasting migraine with time-series machine learning from mobile health data. J. Headache Pain 2026; 27: 91.

12. Holsteen KK, Hittle M, Barad M, Nelson LM. Development and Internal Validation of a Multivariable Prediction Model for Individual Episodic Migraine Attacks Based on Daily Trigger Exposures. Headache 2020; 60: 2364–2379.

13. Houle TT, Turner DP, Golding AN, Porter JAH, Martin VT, Penzien DB, et al. Forecasting Individual Headache Attacks Using Perceived Stress: Development of a Multivariable Prediction Model for Persons With Episodic Migraine. Headache 2017; 57: 1041–1050.

14. Alqassem I, Borole P, Shaker A, Rajan A. Tracing pain: predictive modeling for migraine and headache triggers. In: Proceedings of the 13th IEEE International Conference on Healthcare Informatics (ICHI); 2025.

15. Jongkees BJ, Colzato LS. Spontaneous eye blink rate as predictor of dopamine-related cognitive function—A review. Neurosci. Biobehav. Rev. 2016; 71: 58–82.

16. Katsuki M, Matsumori Y, Kawamura S, Kashiwagi K, Koh A, Tachikawa S, et al. Investigating the effects of weather on headache occurrence using a smartphone application and artificial intelligence: A retrospective observational cross-sectional study. Headache 2023; 63: 585–600.

17. van den Hoek TC, van de Ruit M, Terwindt GM, Tolner EA. EEG Changes in Migraine—Can EEG Help to Monitor Attack Susceptibility? Brain Sci. 2024; 14: 508.

18. Martins IP, Westerfield M, Lopes M, Maruta C, Gil-da-Costa R. Brain state monitoring for the future prediction of migraine attacks. Cephalalgia 2020; 40: 255–265.

19. Shahaf G, Kuperman P, Bloch Y, Yariv S, Granovsky Y. Monitoring Migraine Cycle Dynamics with an Easy-to-Use Electrophysiological Marker—A Pilot Study. Sensors 2018; 18: 3918.

20. Cao Z, Lai KL, Lin CT, Chuang CH, Chou CC, Wang SJ. Exploring resting-state EEG complexity before migraine attacks. Cephalalgia 2018; 38: 1296–1306.

21. De Haan G, Jeanne V. Robust Pulse Rate from Chrominance-Based rPPG. IEEE Trans. Biomed. Eng. 2013; 60: 2878–2886.

22. Verkruysse W, Svaasand LO, Nelson JS. Remote plethysmographic imaging using ambient light. Opt. Express 2008; 16: 21434–21445.

23. Soukupová T, Čech J. Real-time eye blink detection using facial landmarks. In: Proceedings of the 21st Computer Vision Winter Workshop (CVWW); 2016 Feb 3–5; Rimske Toplice, Slovenia.

24. Task Force of the European Society of Cardiology and the North American Society of Pacing and Electrophysiology. Heart rate variability: standards of measurement, physiological interpretation, and clinical use. Circulation 1996; 93: 1043–1065.

25. Ritu, Kumar H, Mehrotra S, Nagappan P. A stability analysis of SHAP and LIME under feature correlation for migraine prediction using deep learning. In: Proceedings of the 4th International Conference on Augmented Intelligence and Sustainable Systems (ICAISS); 2026. p. 753–758.

26. Peduzzi P, Concato J, Kemper E, Holford TR, Feinstein AR. A simulation study of the number of events per variable in logistic regression analysis. J. Clin. Epidemiol. 1996; 49: 1373–1379.

27. Collins GS, Moons KGM, Dhiman P, Riley RD, Beam AL, Van Calster B, et al. TRIPOD+AI Statement: Updated Guidance for Reporting Clinical Prediction Models That Use Regression or Machine Learning Methods. BMJ 2024; 385: e078378.

28. Hosmer DW, Lemeshow S, Sturdivant RX. Applied logistic regression. 3rd ed. Hoboken, NJ: Wiley; 2013.

29. Shaffer F, Ginsberg JP. An Overview of Heart Rate Variability Metrics and Norms. Front. Public Health 2017; 5: 258.

30. Dumkrieger GM, Chiang CC, Zhang P, Minen MT, Cohen F, Hranilovich JA. Artificial Intelligence Terminology, Methodology, and Critical Appraisal: A Primer for Headache Clinicians and Researchers. Headache 2025; 65: 180–190.

31. Buse DC, McGinley JS, Lipton RB. Predicting the Future of Migraine Attack Prediction. Headache 2020; 60: 2125–2128.

32. Provenzi L, Calcaterra V, Nazzari S, Agnelli PO, Xodo M, De Pasquale S, et al. The Many Faces of Stress: Preliminary Validation of a Remote Photoplethysmography-Based Tool for Psychophysiological Stress and Emotional Distress Monitoring. Healthcare 2026; 14: 1893.

33. Dumkrieger GM. The Promise of Machine Learning in Predicting Migraine Attacks. Cephalalgia 2025; 45: 03331024251391207.

